# A Novel Cohorting and Isolation Strategy for Suspected COVID-19 Cases during a Pandemic

**DOI:** 10.1101/2020.05.14.20091843

**Authors:** Benjamin Patterson, Michael Marks, Gema Martinez-Garcia, Gabriella Bidwell, Akish Luintel, Dalia Ludwig, Tom Parks, Philip Gothard, Rik Thomas, Sarah Logan, Karen Shaw, Neil Stone, Mike Brown

**Affiliations:** Department of Clinical Microbiology, Unviersity College London; Hospital for Tropical Diseases, University College London; Clinical Research Department, London School of Hygiene & Tropical Medicine; Division of Infection, University College London; Department of Acute Medicine, University College London; Intensive Care Department, University College London

## Abstract

**Introduction:** The COVID-19 pandemic presents a significant infection prevention and control challenge. The admission of large numbers of patients with suspected COVID-19 disease risks overwhelming the capacity to protect other patients from exposure. The delay between clinical suspicion and confirmatory testing adds to the complexity of the problem.

**Methods:** We implemented a triage tool aimed at minimising hospital acquired COVID-19 particularly to patients at risk of severe disease. Patients were allocated to triage categories defined by likelihood of COVID-19 and risk of a poor outcome. Category A (low-likelihood; high-risk), B (high-likelihood; high-risk), C (high-likelihood; low-risk) and D (low-likelihood; low-risk). This determined the order of priority for isolation in single-occupancy rooms with Category A the highest. Patients in other groups were cohorted when isolation capacity was limited with additional interventions to reduce transmission.

**Results:** 93 patients were evaluated with 79 (85%) receiving a COVID-19 diagnosis during their admission. Of those *without* a COVID-19 diagnosis: 10 were initially triaged to Category A; 0 to B; 1 to C and 4 to D. All high risk patients requiring isolation were, therefore, admitted to single-occupancy rooms and protected from exposure. 28 (30%) suspected COVID-19 patients were evaluated to be low risk (groups C & D) and eligible for cohorting. No symptomatic hospital acquired infections were detected in the cohorted patients.

**Discussion:** Application of a clinical triage tool to guide isolation and cohorting decisions may reduce the risk of hospital acquired transmission of COVID-19 especially to individuals at the greatest of risk of severe disease.

## Background

Since its emergence in December 2019, the COVID-19 pandemic has placed substantial burdens on health systems globally. Avoiding healthcare associated transmission is a major challenge for both primary and secondary care facilities. In the UK, all individuals admitted to hospitals with clinical syndromes of pneumonia, severe acute respiratory infections (SARI) and influenza-like illness (ILI) represent suspected cases of COVID-19 and are eligible for testing according to PHE guidelines [1]. The gold-standard means of COVID-19 diagnosis remains molecular laboratory testing by PCR of a nasopharyngeal swab. In most UK hospitals this test is performed in a laboratory, with a turnaround time of several hours if not longer, and so patients are received to wards with unconfirmed COVID-19 status at time of admission. One aspect of standard infection prevention and control (IPC) practice is to place a patient with suspicion of a transmissible infection into a single-occupancy room with appropriate IPC precautions pending the results of investigations. This protects other patients and staff from potential transmission. However, the high burden of COVID-19 cases has overwhelmed the availability of single-occupancy rooms in many hospitals.

In the absence of a near patient rapid diagnostic test, clinicians need a tool to assess the probability of COVID-19 on initial assessment, and triage based on epidemiological risk factors, routine investigations and bedside observations. This enables safe isolation or appropriate cohorting of suspected COVID-19 cases. A particular challenge is presented with COVID-19, as this is a new disease entity and consequently there is limited experience of the clinical, radiological and laboratory features. However, early data and case series have highlighted a number of clinical features which may be of value in identifying patients with COVID-19 amongst patients presenting to hospital with other causes of pneumonia, SARI and ILI. Suggested markers include lymphopenia [2], bilateral chest x-ray infiltrates [2] and absence of neutrophilia. A weakness of current data is that many of these markers are non-specific and there are no high-quality studies describing how accurately they differentiate COVID-19 from other causes of pneumonia, SARI or ILI.

Therefore, there is an urgent need to evaluate the performance of these proposed markers in a real world setting. Specifically their predictive value to optimise allocation of single-occupancy rooms to shield the most vulnerable patients. Within the context of the COVID-19 pandemic, we evaluated a novel triaging tool based on both clinical probability of infection and an individuals’ comorbidities at a large tertiary referral centre in London, UK.

## Methods

At University College London Hospital (UCLH), we implemented a system of triaging patients on the basis of clinical features suggestive of COVID-19, age and comorbidities. All admitted patients who met the case definition for COVID-19 testing were allocated to one of four categories in the Emergency Department (see Figure 1).

**Figure 1.**
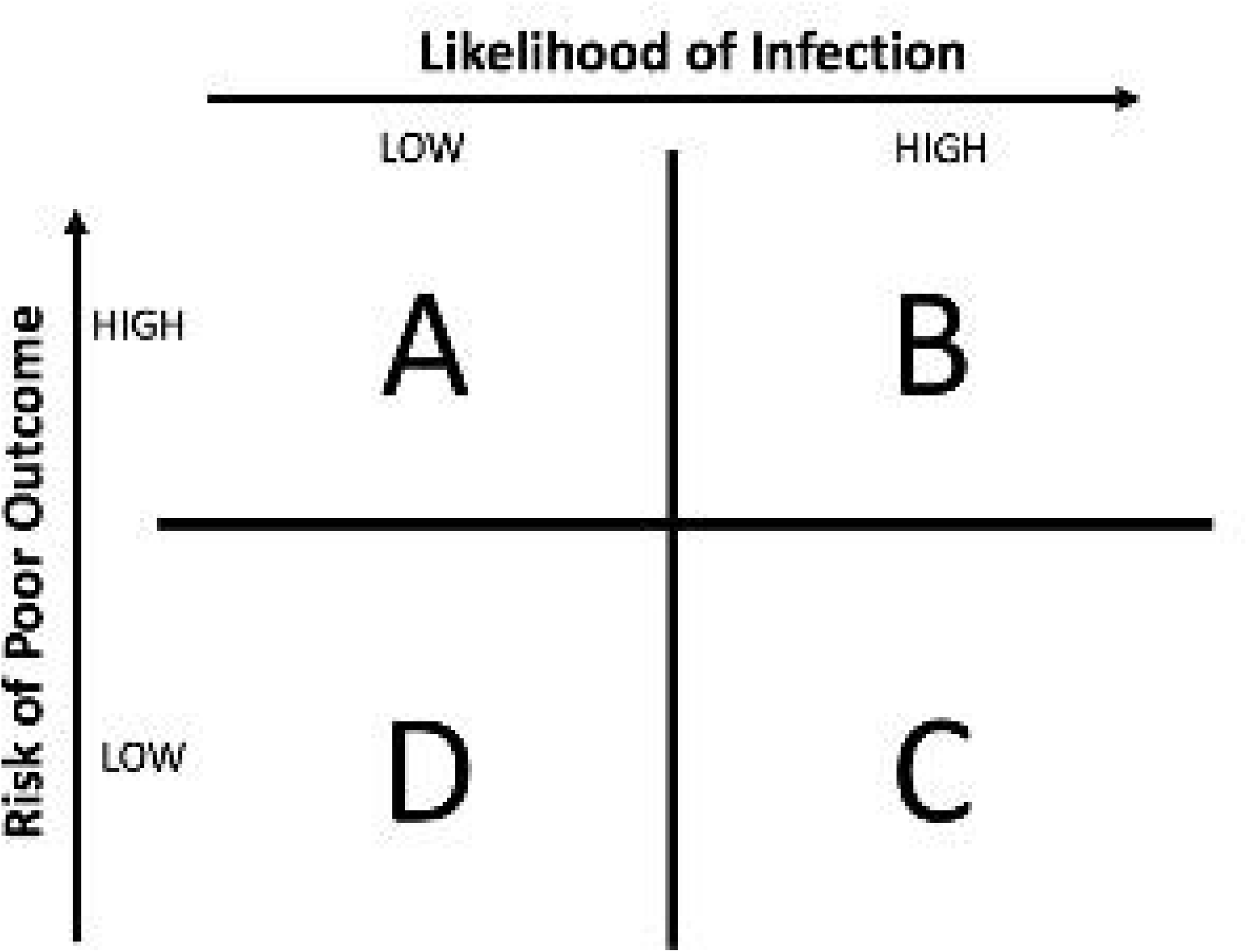
Simple 2×2 table illustrating the characteristics of the four triage categories.

Category A represented patients clinically evaluated to have a low likelihood of COVID-19 but significant comorbidities. These patients were given the highest priority for single-occupancy rooms. Category B and C represented patients considered to have a high probability of COVID-19, with category B patients having significant comorbidities. Category B patients were therefore second priority for single-occupancy rooms. When such rooms were unavailable these patients were cohorted in reduced occupancy multi-bedded bays on wards designated for suspected COVID-19 patients. Category C patients had minimal comorbidities thus were typically cohorted in the same reduced occupancy multi-bedded bays with other category C or B patients. This maintained availability of single-occupancy rooms for category A patients. Finally, category D patients were considered to be both a low clinical probability of COVID-19 and without significant comorbidities. These individuals were cohorted together on the same ward as suspected COVID-19 patients in a designated low probability bay.

To implement this strategy an infectious diseases clinician stationed in the emergency department applied an isolation and cohorting algorithm (see figure 2). This clinician assisted the admitting medical team in assessing the clinical probability of COVID-19. This assessment combined clinical skills with investigation results and evaluation of the extent of the comorbidities. On occasion, discussion with a radiologist aided the decision making process. Priority for single-occupancy rooms was determined through the triage category allocation.

**Figure 2.**
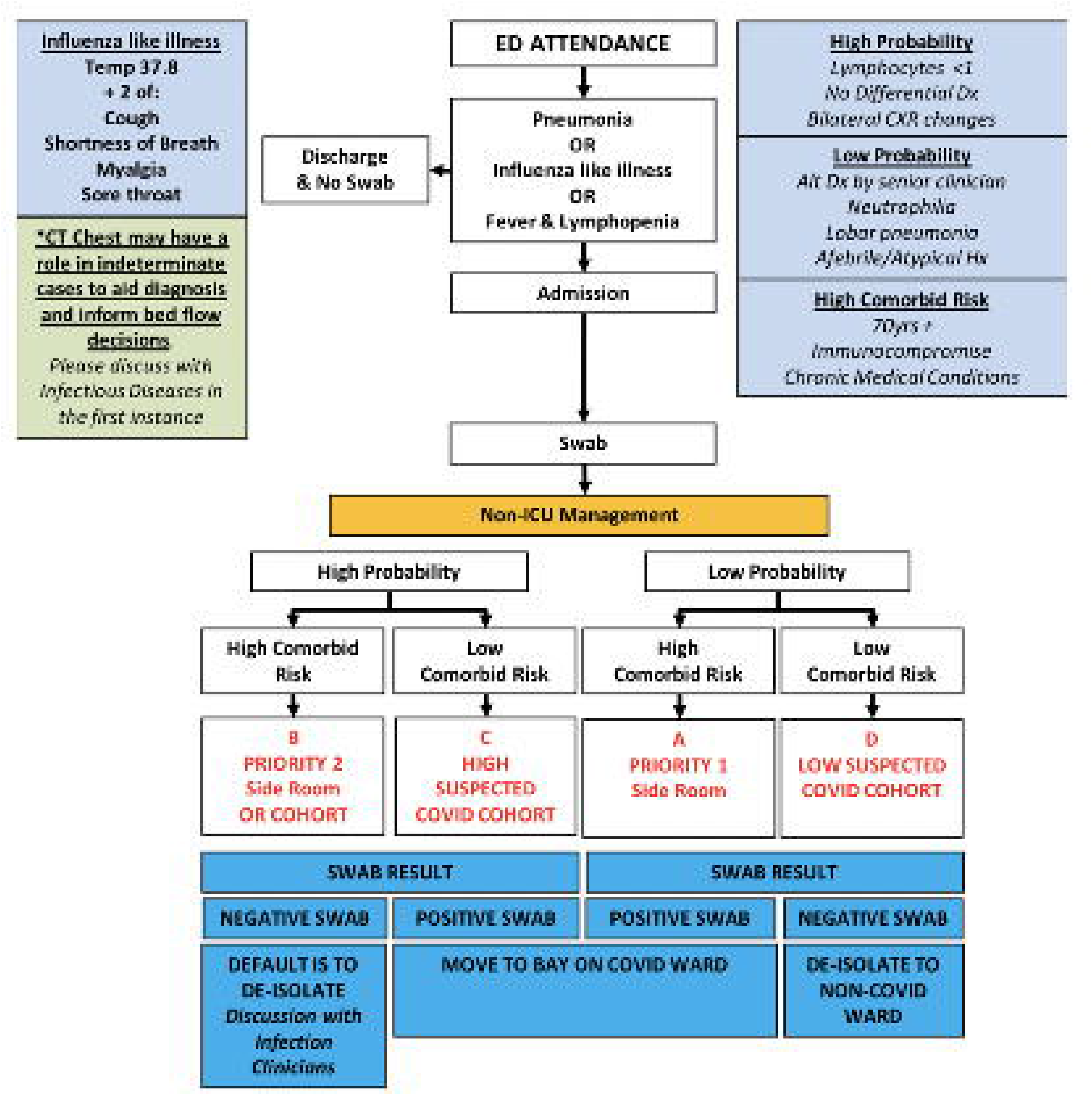
Isolation and cohorting algorithm

We collected data on the categorisation assigned by the infectious diseases clinician, clinical information available at the time of presentation and whether an eventual diagnosis of COVID-19 was confirmed based on clinical, radiological and molecular criteria. Here we report results of an initial evaluation of this triage tool.

Statistical analyses were performed using the R statistical program (R Core Team 2019). Comparisons were made using the non-parametric Wilcoxon rank sum tests for continuous variables and Fisher’s exact tests for discrete variables.

Ethics: This study was assessed through the NHS Health Research Authority system (HRA) (http://www.hra-decisiontools.org.uk/research/) and by the Research and Audit committee of the Hospital for Tropical Diseases and was found to meets the UK NHS definition of a retrospective service evaluation for which formal ethical review was therefore not required

## Results

99 patients suspected of having COVID-19 were admitted to UCLH between 27th March and 2nd April 2020. 93 were prospectively given a triage category and had a subsequent nasopharyngeal swab result (SARS-CoV 2 RT-PCR) available.

Figure 3 highlights the proportion of those admitted with suspicion of COVID-19 who were allocated to each category by clinician assessment of the clinical variables. 60% (15 out of 25) of individuals in Category A were eventually diagnosed with COVID-19; 100% (40 out of 40) in category B; 96% (23 out of 24) in category C and 25% (1 out of 4) in category D. 28 (30%) patients in categories C and D were therefore considered appropriate for cohorting in the designated COVID-19 suspect ward with low risk of poor outcome in event of a hospital transmission.

**Figure 3.**
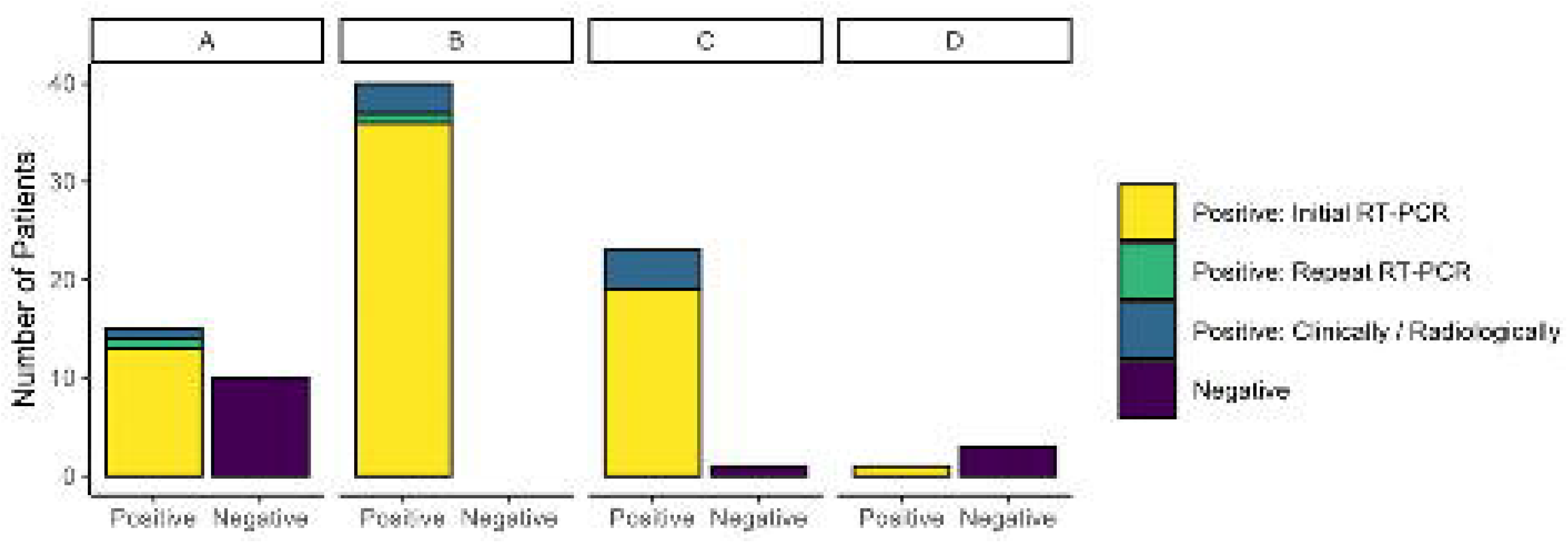
Number of patients allocated to each triage category divided by eventual diagnosis.

Comparison between prediction of COVID-19 and PCR confirmation by nasopharyngeal swab was made by combining the triage categories (B&C) predicting a high probability and those (A&D) predicting a low probability. A significant association was found between prediction and swab result (chi squared statistic 13.2; p<0.001).

68 (73%) patients were found to have a positive swab. Of the 25 negative swabs, 11 (44%) were re-evaluated by the infectious diseases team after the result and diagnosed as highly likely to be COVID-19 based on review of radiology and clinical information. In these cases the swab result was considered to be negative most likely because of the advanced stage of the illness and the known decline in nasopharyngeal RNA yield at this stage [3]. In two of these cases a repeat nasopharyngeal swab was positive. These cases either remained in single-occupancy rooms or were transferred to COVID-19 positive cohort wards throughout their admission. Of the 14 COVID-19 negative cases, 10 were triaged to category A, 0 to category B, 1 to category C and 3 to category D. Those in category A were initially admitted to single-occupancy rooms and for the 3 patients in category D admitted to COVID-19 cohort bays, only one individual was exposed to positive cases. For this individual no symptoms of COVID-19 developed within the 14 days to indicate a healthcare associated transmission (see Table 1).

**Table 1.** Admission locations and exposures for all COVID-19 negative individuals.

| **Triage Category** | **Number of Cases** | **Admission Location** | **Exposure to COVID-19 patients (within the same bay)** |
| --- | --- | --- | --- |
| A | 10 | All Single-Occupancy Rooms | None |
| B | 0 | N/A | N/A |
| C | 1 | Single-Occupancy Room | None |
| D | 3 | COVID Suspect Bays<br>(reserved for Group D) | 1 patient exposed* |

\*No symptoms of infection in the 14 days following exposure.

Table 2 illustrates the differences in presenting features used to determine high and low probability of COVID-19. Patients assigned to the high likelihood triage categories typically had a longer illness duration and were more likely to present with a cough and/or fever. Oxygen requirement at presentation was significantly higher in the high probability group and the chest imaging more likely to show bilateral disease. Table 3 demonstrates differences in age, comorbidities and the Rockwood Frailty Score [4] used to determine high risk and low risk groups. Patients assigned to the high risk groups were significantly older and more frequently had cardiovascular and cerebrovascular comorbidities in line with recent reporting [5].

**Table 2.** Comparison of presenting symptoms between the triage categories with patients assessed as high likelihood (B & C) and low likelihood (A & D) of COVID-19.

|  | High Likelihood<br>(B & C) | Low Likelihood<br>(A & D) | p-value |
| --- | --- | --- | --- |
| n | 64 | 29 |  |
| **Illness Duration** |  |  |  |
| median days [IQR] | 7 [4, 10] | 4 [2, 7.5] | 0.050 |
| **Symptoms** |  |  |  |
| Cough (%) | 41 (64.1) | 11 (37.9) | 0.034 |
| Shortness of Breath (%) | 28 (43.8) | 13 (44.8) | >0.99 |
| Fevers (%) | 38 (59.4) | 9 (31.0) | 0.021 |
| Myalgia (%) | 8 (12.5) | 1 (3.4) | 0.323 |
| Gastrointestinal<br>Symptoms (%) | 4 (6.2) | 2 (6.9) | >0.99 |
| Other Symptoms* (%) | 9 (14.1) | 7 (24.1) | 0.370 |
| **Oxygen Requirement (%)** |  |  | 0.010 |
| No Oxygen Required | 15 (23.4) | 16 (55.2) |  |
| Nasal Cannula (1-4L) | 15 (23.4) | 5 (17.2) |  |
| Higher Oxygen Requirement | 34 (53.1) | 8 (27.6) |  |
| **Laboratory Values** |  |  |  |
| Neutrophilia ($>7.5 \times 10^9/L$) (%) | 18 (28.1) | 13 (46.4) | 0.142 |
| Lymphopenia ($<1.2 \times 10^9/L$) (%) | 44 (68.8) | 14 (50.0) | 0.139 |
| **Chest Radiography (%)** |  |  | **<0.001** |
| Bilateral Infiltrates | 43 (67.2) | 7 (24.1) |  |
| Unilateral Infiltrates | 12 (18.8) | 2 (6.9) |  |
| Indeterminant | 0 (0.0) | 4 (13.8) |  |
| No Chest X-ray Changes | 9 (14.1) | 16 (55.2) |  |

\*Other Symptoms include: sore throat, wheeze, confusion, fall

**Table 3.**
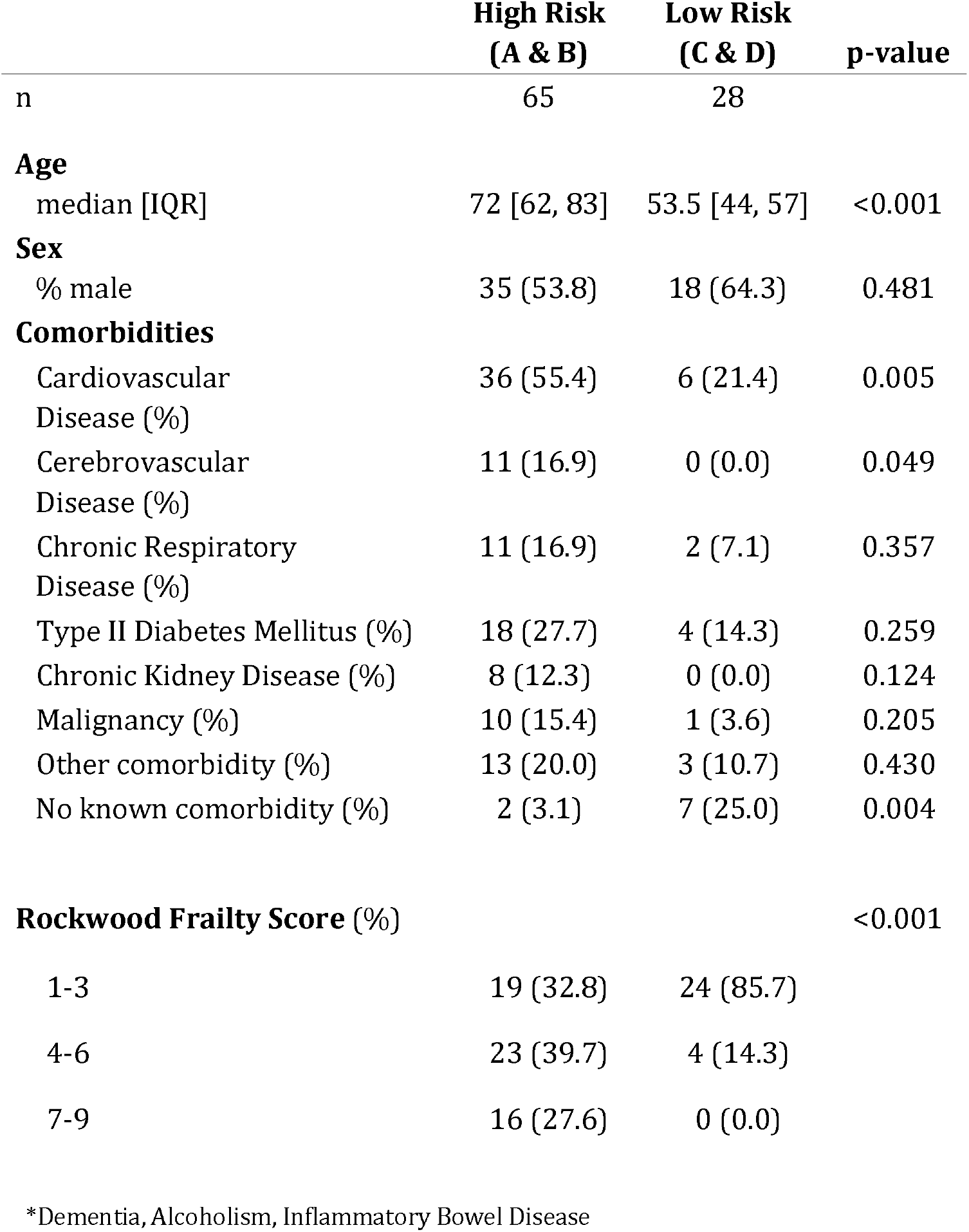
Comparison of age, pre-existing patient comorbidities and Rockwood Frailty Score between triage categories with patients assessed as high risk (A & B) and low risk (C & D) of a poor outcome from COVID-19.

|  | High Risk<br>(A & B) | Low Risk<br>(C & D) | p-value |
| --- | --- | --- | --- |
| n | 65 | 28 |  |
| **Age** |  |  |  |
| median [IQR] | 72 [62, 83] | 53.5 [44, 57] | <0.001 |
| **Sex** |  |  |  |
| % male | 35 (53.8) | 18 (64.3) | 0.481 |
| **Comorbidities** |  |  |  |
| Cardiovascular Disease (%) | 36 (55.4) | 6 (21.4) | 0.005 |
| Cerebrovascular Disease (%) | 11 (16.9) | 0 (0.0) | 0.049 |
| Chronic Respiratory Disease (%) | 11 (16.9) | 2 (7.1) | 0.357 |
| Type II Diabetes Mellitus (%) | 18 (27.7) | 4 (14.3) | 0.259 |
| Chronic Kidney Disease (%) | 8 (12.3) | 0 (0.0) | 0.124 |
| Malignancy (%) | 10 (15.4) | 1 (3.6) | 0.205 |
| Other comorbidity (%) | 13 (20.0) | 3 (10.7) | 0.430 |
| No known comorbidity (%) | 2 (3.1) | 7 (25.0) | 0.004 |
| **Rockwood Frailty Score (%)** |  |  | **<0.001** |

|  |  |  |
| --- | --- | --- |
| 1-3 | 19 (32.8) | 24 (85.7) |
| 4-6 | 23 (39.7) | 4 (14.3) |
| 7-9 | 16 (27.6) | 0 (0.0) |

\*Dementia, Alcoholism, Inflammatory Bowel Disease

## Discussion

This study evaluated the use of a pragmatic triage tool for prompt isolation or cohorting of patients in the context of the COVID-19 pandemic. The tool was designed to manage patient flow, in the event of insufficient single-occupancy rooms to isolate all suspected cases at admission. The focus was to prevent healthcare associated transmission and, in particular, to identify and protect individuals at the greatest risk of a poor outcome should a new infection occur. We therefore focused not merely on the risk of COVID-19, but on the risk to patients of misclassification. To this end we found the use of well-described clinical, laboratory and radiological markers as predictors of molecularly confirmed COVID-19 disease to have high positive predictive value. Partial or total absence of typical features did not rule out COVID-19 disease, but allowed us to identify a subgroup of patients with a higher likelihood of a diagnosis other than COVID-19 for whom the single-occupancy rooms could be reserved.

Given the volume of acute admissions with suspicion for COVID-19 disease, application of the usual process of isolation for each patient with COVID-19 would have overwhelmed the single-occupancy room capacity. By implementing this system we were able to ensure the most vulnerable individuals admitted during this five day period were correctly prioritised for single-occupancy rooms. Importantly, the group, characterised as low probability of disease but with high comorbidities (Group A), identified ten cases who were ultimately deemed to be negative for COVID-19. These individuals were effectively shielded from SARS-CoV-2 exposure and risk of healthcare associated transmission during their admission. At the same time all but one of the high probability cases were diagnosed with COVID-19 based on nasopharyngeal swab, clinical and radiological criteria. Many of these patients were therefore appropriately cohorted without incurring additional risk related to exposure, but with reduction in bed pressure for single-occupancy rooms.

Key to the implementation of this triage tool was the creation of confirmed and suspected COVID-19 wards areas. Capacity for this was facilitated by cancellation of all elective services. These ward areas were physically separated by constructing doors, with a one-way flow of staff entering and exiting the ward. Personal protective equipment (PPE) donning and doffing stations were positioned at these fixed points of entry and exit. In order to maintain strict separation, both patient and staff pathways were redesigned necessitating the closure of communal staff areas. Bed spacing within bays was expanded by removing beds to increase the distance between patients, and all non-essential equipment was removed. Bedside equipment was not shared between bays to reduce the extent of environmental contamination. Category D patients (low likelihood of infection) were assigned specific bays to minimise their physical proximity to patients in other groups and were placed furthest from the doffing station in the least contaminated areas of the COVID-19 suspect ward. Aerosol generating procedures were avoided unless in single-occupancy rooms. For all patients, once the nasopharyngeal swab result was available relocation of patients was determined collaboratively by the IPC team and infection clinicians.

A limitation of this evaluation includes the specificity to the contemporaneous COVID-19 prevalence. From late March to early April, the UK saw a very high rate of cases admitted to hospitals in London and very likely a reduced attendance of individuals with other medical problems. Therefore the pre-test probability of COVID-19 was extremely high which may have impacted our results. Secondly the triage was conducted by an experienced infectious diseases clinician. The extent to which the accuracy of the triage can be generalised may depend on the identification of a combination of objective markers with adequate predictive value in a range of settings.

## Conclusion

In summary, our evaluation demonstrated that early assessment of patients with suspected COVID-19, by a clinician with appropriate expertise, effectively identified a high risk cohort most appropriate for isolation. This approach combined with innovative IPC measures reduced bed pressures without increasing the risk of healthcare associated transmission. This triage tool may be of value more generally in health systems responding to the ongoing COVID-19 pandemic, particularly during sustained transmission of the virus when pre-test probability of COVID-19 positivity is high.

## Data Availability

All relevant data is contained in the manuscript

